# High-throughput sequencing of SARS-CoV-2 in wastewater provides insights into circulating variants

**DOI:** 10.1101/2021.01.22.21250320

**Authors:** Rafaela S. Fontenele, Simona Kraberger, James Hadfield, Erin M. Driver, Devin Bowes, LaRinda A. Holland, Temitope O.C. Faleye, Sangeet Adhikari, Rahul Kumar, Rosa Inchausti, Wydale K. Holmes, Stephanie Deitrick, Philip Brown, Darrell Duty, Ted Smith, Aruni Bhatnagar, Ray A. Yeager, Rochelle H. Holm, Natalia Hoogesteijn von Reitzenstein, Elliott Wheeler, Kevin Dixon, Tim Constantine, Melissa A. Wilson, Efrem S. Lim, Xiaofang Jiang, Rolf U. Halden, Matthew Scotch, Arvind Varsani

**Author notes:** Corresponding authors Rafaela S. Fontenele, Simona Kraberger, Arvind Varsani. Authors contributed equally to this work (Rafaela S. Fontenele, Simona Kraberger).

## Abstract

Severe acute respiratory syndrome coronavirus 2 (SARS-CoV-2) emerged from a zoonotic spill-over event and has led to a global pandemic. The public health response has been predominantly informed by surveillance of symptomatic individuals and contact tracing, with quarantine, and other preventive measures have then been applied to mitigate further spread. Non-traditional methods of surveillance such as genomic epidemiology and wastewater-based epidemiology (WBE) have also been leveraged during this pandemic. Genomic epidemiology uses high-throughput sequencing of SARS-CoV-2 genomes to inform local and international transmission events, as well as the diversity of circulating variants. WBE uses wastewater to analyse community spread, as it is known that SARS-CoV-2 is shed through bodily excretions. Since both symptomatic and asymptomatic individuals contribute to wastewater inputs, we hypothesized that the resultant pooled sample of population-wide excreta can provide a more comprehensive picture of SARS-CoV-2 genomic diversity circulating in a community than clinical testing and sequencing alone. In this study, we analysed 91 wastewater samples from 11 states in the USA, where the majority of samples represent Maricopa County, Arizona (USA). With the objective of assessing the viral diversity at a population scale, we undertook a single-nucleotide variant (SNV) analysis on data from 52 samples with >90% SARS-CoV-2 genome coverage of sequence reads, and compared these SNVs with those detected in genomes sequenced from clinical patients. We identified 7973 SNVs, of which 5680 were “novel” SNVs that had not yet been identified in the global clinical-derived data as of 17^th^ June 2020 (the day after our last wastewater sampling date). However, between 17^th^ of June 2020 and 20^th^ November 2020, almost half of the SNVs have since been detected in clinical-derived data. Using the combination of SNVs present in each sample, we identified the more probable lineages present in that sample and compared them to lineages observed in North America prior to our sampling dates. The wastewater-derived SARS-CoV-2 sequence data indicates there were more lineages circulating across the sampled communities than represented in the clinical-derived data. Principal coordinate analyses identified patterns in population structure based on genetic variation within the sequenced samples, with clear trends associated with increased diversity likely due to a higher number of infected individuals relative to the sampling dates. We demonstrate that genetic correlation analysis combined with SNVs analysis using wastewater sampling can provide a comprehensive snapshot of the SARS-CoV-2 genetic population structure circulating within a community, which might not be observed if relying solely on clinical cases.

## 1. Introduction

Severe acute respiratory syndrome coronavirus 2 (SARS-CoV-2) is the bigest pandemic since the 1918 H1N1 influenza A virus (Wang et al., 2020; Yan et al., 2020). The SARS-CoV-2 outbreak in humans likely emerged from a zoonotic transmission event(s), and was first recorded in December, 2019, in the City of Wuhan, China (Andersen et al., 2020; Boni et al., 2020; Zhang and Holmes, 2020). According to the Johns Hopkins Coronavirus Resource Center (Dong et al., 2020), there have been >95 million confirmed cases, resulting in more than >2 million deaths globally as of 18^th^ January 2021. SARS-CoV-2 is a positive-sense single-stranded RNA virus in the family *Coronaviridae* (Gorbalenya et al., 2020) that can cause a range of symptoms in infected individuals including complications with breathing, dry cough, fever, and diarrhoea (Wang et al., 2020). However, the majority of individuals show little to no symptoms (Buitrago-Garcia et al., 2020; Byambasuren et al., 2020; Kimball et al., 2020; Syangtan et al., 2020).

Clinical testing of individuals for SARS-CoV-2 is the primary surveillance method for informing public health strategic interventions, and essential for implementing preventive measures, such as quarantine, to mitigate the spread of the virus. The most frequently used approach for clinical testing relies on the detection of genomic elements of SARS-CoV-2 by reverse transcription-quantitative polymerase chain reaction (RT-qPCR) based methods (CDC, 2020a; WHO). The clinical analysis is now also being complemented with antibody-based assays (Adams et al., 2020; Becker et al., 2020; Bryant et al., 2020; CDC, 2020b; WHO) that provide an indication of current or previous exposure to SARS-CoV-2.

High-throughput sequencing (HTS) technologies are being used to sequence the SARS-CoV-2 genome from a subset of the infected population globally using clinical samples. This has resulted in over >278,000 published genomes (Elbe and Buckland-Merrett, 2017; Shu and McCauley, 2017), and has provided insight into its origins, spread, and diversity via computational approaches in genomic epidemiology. Screening/testing of a large number of individuals for SARS-CoV-2 can be challenging particularly from a logistics perspective due to sample collection and transportation, availability and storage of assay reagents, and the rapid turnaround time needed for test results to be most informative to healthcare outcomes and pandemic management. Furthermore, in most countries it is largely the symptomatic population that is targeted for testing and therefore a large proportion of infected asymptomatic individuals may be missed.

Nasopharyngeal swabs and saliva samples have been the principal sample types used for screening; however, SARS-CoV-2 has also been detected in other clinical specimens such as faeces, from both symptomatic and asymptomatic infected individuals (Chen et al., 2020; Jones et al., 2020; Park et al., 2020; Tang et al., 2020; Xing et al., 2020). Moreover, of late, wastewater samples have been utilized as a way to identify several pathogenic human viruses and, not surprisingly, it has gained attention for assessing population-level trends of SARS-CoV-2 infections.

Detection of SARS-CoV-2 in wastewater (untreated and treated) has been a focus of research, with feasibility highlighted in the review by Farkas et al. (2020) and with reported studies from locations including North America (D’Aoust et al., 2021; Nemudryi et al., 2020; Peccia et al., 2020; Wu et al., 2020), Europe (Balboa et al., 2020; Kocamemi et al., 2020; La Rosa et al., 2020; Medema et al., 2020; Randazzo et al., 2020; Westhaus et al., 2021; Wurtzer et al., 2020), Asia (Kumar et al., 2020; Zhang et al., 2020) and Oceania (Ahmed et al., 2020). These studies used a range of sample concentration and viral RNA recovery approaches followed by RT-qPCR amplification to detect and determine the viral load. These proof of concept studies demonstrated the detection of SARS-CoV-2 in wastewater and identified trends indicating wastewater monitoring can serve as a useful early warning tool for informing public health (Farkas et al., 2020). Although some studies did verify, by sequencing, the RT-qPCR products were indeed detecting SARS-CoV-2, most rely on the threshold cycle (Ct) values of RT-qPCR assays. Beyond this, two recent studies have sequenced the SARS-CoV-2 genomes recovered from wastewater (Crits-Christoph et al., 2021; Izquierdo Lara et al., 2020).

Despite the promising success of these prior studies, it is still unclear how well wastewater-based epidemiology can identify the genetic diversity of SARS-CoV-2 in a given population and how this relates to known viral diversity of clinical cases. This is especially important as new lineages are being discovered. For example, the B.1.351 strain in the United Kingdom that contains single-nucleotide variants (SNVs) of potential biological significance such as N501Y (in the spike protein) (Rambaut et al., 2020b) and K417N, E484K and N501Y in South Africa (Tegally et al., 2020). To investigate the potential of using wastewater to gain insights into variants of SARS-CoV-2 circulating in the population, we used a tiling amplicon-based high-throughput sequencing approach to determine SARS-CoV-2 sequences (spanning the genome) in 91 wastewater samples collected from 11 states in the United States (USA) between 7^th^ April 2020 and 16^th^ June 2020. To further survey the viral diversity circulating within a community and to examine how these relate to sequences from clinical cases, we undertook SNV analysis and beta diversity analyses of SARS-CoV-2 sequences in 52 (>90% coverage) out of the 91 wastewater samples from 10 states. We focused specifically on spatial and temporal trends, and how they compare with clinically-derived data.

## 2. Material and methods

### 2.1. Sample collection and transport

Flow- or time-weighted, 24-hr composite samples of untreated wastewater were collected either from the headworks of the wastewater treatment plant, from within the wastewater collection system or at hospital facilities using high frequency automated samplers (Teledyne ISCO, USA) from locations across 11 states in the USA between 7^th^ April 2020 and 16^th^ June 2020 (Table 1, Figure 1A, Sup Figure 1). Most samplers had refrigeration capabilities or were supplied with an ice/dry ice blend to keep the interior collection vessel cool. During sample collection, wastewater was thoroughly mixed and transferred to high-density polyethylene sample bottles and placed on ice for transport. The samples were either hand delivered or shipped (next-day/2-day) in insulated shipping containers for subsequent processing and analysis.

**Table 1:** Summary of wastewater sample information. The collection date reflects influent from the previous day. Details of the location including state, city, and region of collection, and Ct value from the RT-qPCR SARS-CoV-2 detection assay targeting the E gene. The SARS-CoV2 genome percentage coverage based on the HTS for each sample is provided.

| State | Location ID | Sampling date | Sample ID | Ct value | Mean coverage | Percentage coverage | Total reads |
| --- | --- | --- | --- | --- | --- | --- | --- |
| Arizona | G2 | 7-May-20 | 122 | 35.1 | 21.9801 | 37.91 | 8228 |
| Arizona | G2 | 10-Jun-20 | G3 | 32.2 | 82.9204 | 95.7246 | 30944 |
| Arizona | Guadalupe | 6-May-20 | 110 | 31.9 | 139.084 | 97.8267 | 51936 |
| Arizona | Guadalupe | 10-May-20 | 136 | 30.8 | 249.107 | 98.6426 | 93131 |
| Arizona | Guadalupe | 12-May-20 | 147 | 30.2 | 682.605 | 99.0555 | 254395 |
| Arizona | Guadalupe | 16-May-20 | 177 | 30.2 | 800.327 | 98.9946 | 298388 |
| Arizona | Guadalupe | 19-May-20 | 179 | 30.9 | 780.958 | 99.0217 | 291504 |
| Arizona | Guadalupe | 21-May-20 | 203 | 29.9 | 1496.09 | 99.1029 | 558227 |
| Arizona | Guadalupe | 26-May-20 | 227 | 30.6 | 563.257 | 98.9269 | 209969 |
| Arizona | Guadalupe | 30-May-20 | 253 | 28.9 | 1784.29 | 99.1097 | 665406 |
| Arizona | Guadalupe | 3-Jun-20 | 277 | 30.2 | 31.7447 | 71.6733 | 11859 |
| Arizona | Guadalupe | 5-Jun-20 | 303 | 30.6 | 18.0822 | 65.1061 | 6766 |
| Arizona | Guadalupe | 7-Jun-20 | 321 | 30.8 | 457.993 | 98.9269 | 170607 |
| Arizona | Guadalupe | 9-Jun-20 | 341 | 30.8 | 1111.99 | 98.998 | 414806 |
| Arizona | Guadalupe | 11-Jun-20 | 359 | 29.5 | 45.4204 | 83.8868 | 16957 |
| Arizona | M1 | 27-Apr-20 | 80 | 32.7 | 20.8707 | 43.5666 | 7802 |
| Arizona | M1 | 7-May-20 | 117 | 34.9 | 2.24021 | 7.66054 | 880 |
| Arizona | M1 | 26-May-20 | 225 | 35.9 | 13.4329 | 37.7272 | 5035 |
| Arizona | Rural | 24-Oct-19 | R19 | NA | 10.9956 | 1.29989 | 2698 |
| Arizona | Rural | 16-May-20 | 167 | 35.7 | 29.7984 | 54.0537 | 11099 |
| Arizona | Rural | 3-Jun-20 | 269 | 34.4 | 170.102 | 97.0279 | 63422 |
| Arizona | Rural | 6-Jun-20 | 305 | 33.3 | 87.2427 | 96.7435 | 32575 |
| Arizona | Rural | 9-Jun-20 | 338 | 33 | 81.784 | 97.1497 | 30496 |
| Arizona | Rural | 11-Jun-20 | 349 | 31.6 | 81.6799 | 96.0157 | 30520 |
| Arizona | TP01 | 7-Apr-20 | 4 | 35 | 59.1029 | 66.643 | 22076 |
| Arizona | TP01 | 8-Apr-20 | 3 | 37 | 0.646356 | 1.56054 | 255 |
| Arizona | TP01 | 17-Apr-20 | 57 | 35 | 4.45655 | 15.1958 | 1667 |
| Arizona | TP01 | 21-Apr-20 | 59 | 33 | 18.1784 | 39.5586 | 6761 |
| Arizona | TP01 | 29-Apr-20 | 93 | 35 | 11.943 | 38.2418 | 4446 |
| Arizona | TP01 | 12-May-20 | 137 | 34.7 | 47.4554 | 62.7061 | 17703 |
| Arizona | TP01 | 26-May-20 | 220 | 35.5 | 35.8432 | 64.4122 | 13421 |
| Arizona | TP01 | 2-Jun-20 | 260 | 33.6 | 586.011 | 99.0183 | 218520 |
| Arizona | TP01 | 7-Jun-20 | 322 | 35.7 | 39.971 | 77.0048 | 14903 |
| Arizona | TP01 | 9-Jun-20 | 348 | 31.5 | 339.292 | 98.9066 | 126569 |
| Arizona | TP02 | 29-Apr-20 | 94 | 35 | 2.23134 | 7.12907 | 844 |
| Arizona | TP02 | 12-May-20 | 138 | 35.8 | 5.71064 | 11.9055 | 2144 |
| Arizona | TP02 | 30-May-20 | 247 | 35.1 | 52.7047 | 91.0226 | 19682 |
| Arizona | TP02 | 2-Jun-20 | 261 | 32.6 | 106.321 | 96.0699 | 39581 |
| Arizona | TP02 | 5-Jun-20 | 299 | 34 | 84.0252 | 96.3779 | 31348 |
| Arizona | TP02 | 9-Jun-20 | 344 | 32.8 | 258.612 | 99.1165 | 96441 |
| Arizona | TP03 | 6-Jun-20 | 312 | 34.5 | 130.712 | 97.2344 | 48711 |
| Arizona | TP03 | 7-Jun-20 | 323 | 35.4 | 151.054 | 98.3514 | 56337 |
| Arizona | TP04 | 28-May-20 | 274 | 34.5 | 34.992 | 71.6699 | 13061 |
| Arizona | TP04 | 4-Jun-20 | 288 | 33 | 110.474 | 96.2053 | 41202 |
| Arizona | TP04 | 5-Jun-20 | 129 | 32.7 | 31.8066 | 72.3368 | 11897 |
| Arizona | TP04 | 6-Jun-20 | 314 | 34.7 | 191.419 | 98.8829 | 71379 |
| Arizona | TP04 | 8-Jun-20 | 336 | 32.8 | 220.449 | 98.9371 | 82296 |
| Arizona | TP05 | 25-Apr-20 | 69 | 31.2 | 15.223 | 41.1699 | 5678 |
| Arizona | TP05 | 7-May-20 | 118 | 32.1 | 22.2285 | 50.7803 | 8291 |
| Arizona | TP05 | 19-May-20 | 181 | 35.8 | 38.4298 | 59.3514 | 14304 |
| Arizona | TP05 | 7-Jun-20 | 326 | 35.6 | 27.9763 | 66.1792 | 10443 |
| Arizona | TP05 | 9-Jun-20 | 347 | 26.8 | 3735.92 | 99.1097 | 1510084 |
| Arizona | TP05 | 11-Jun-20 | 358 | 31.5 | 37.94 | 75.9453 | 14211 |
| Arizona | TP06 | 26-Apr-20 | 78 | 34.9 | 2.9937 | 5.98152 | 1127 |
| Arizona | TP06 | 21-May-20 | 198 | 34.9 | 17.187 | 57.3034 | 6445 |
| Arizona | TP06 | 28-May-20 | 234 | 34.7 | 784.976 | 98.998 | 292585 |
| Arizona | TP06 | 3-Jun-20 | 271 | 33.3 | 61.7264 | 93.4159 | 23022 |
| Arizona | TP06 | 5-Jun-20 | 296 | 32.8 | 92.836 | 97.3901 | 34617 |
| Arizona | TP06 | 7-Jun-20 | 318 | 34.6 | 40.5103 | 90.8805 | 15096 |
| Arizona | TP06 | 9-Jun-20 | 339 | 32.6 | 33.4383 | 86.1074 | 12474 |
| Arizona | TP06 | 11-Jun-20 | 351 | 30.6 | 20.0344 | 65.7696 | 7472 |
| Colorado | CO1 | 20-May-20 | Jac_51 | 32.1 | 85.5393 | 93.1282 | 31953 |
| Colorado | CO1 | 28-May-20 | Jac_103 | 34 | 91.4798 | 96.124 | 34120 |
| Georgia | GA1 | 14-May-20 | Jac_33 | 29 | 68.8532 | 94.4078 | 25686 |
| Idaho | ID1 | 18-May-20 | Jac_56 | 34.7 | 88.5662 | 91.114 | 33005 |
| Idaho | ID1 | 25-May-20 | Jac_87 | 35.3 | 113.577 | 94.4416 | 42320 |
| Illinois | IL1 | 19-May-20 | Jac_45 | 33.3 | 79.0705 | 96.9331 | 29490 |
| Illinois | IL1 | 1-Jun-20 | Jac_106 | 33.1 | 54.4429 | 85.8332 | 20365 |
| Illinois | IL2 | 7-May-20 | Jac_12 | 33 | 71.8524 | 90.6875 | 26744 |
| Illinois | IL2 | 1-Jun-20 | Jac_127 | 31.9 | 77.5081 | 87.7357 | 28850 |
| Kansas | KA1 | 20-May-20 | Jac_58 | 33.2 | 91.4503 | 91.9265 | 34117 |
| Kansas | KA1 | 27-May-20 | Jac_96 | 31.7 | 364.619 | 98.9845 | 135932 |
| Kentucky | S1 | 23-Apr-20 | Lou_2 | 33.8 | 31.4104 | 70.7017 | 11723 |
| Kentucky | S2 | 9-Jun-20 | Lou_40 | 33.8 | 352.012 | 98.734 | 131165 |
| Kentucky | S3 | 21-May-20 | Lou_15 | 35.3 | 11.7138 | 36.1193 | 4379 |
| Kentucky | S3 | 28-May-20 | Lou_23 | 35.5 | 9.75725 | 33.6448 | 3640 |
| Kentucky | S3 | 9-Jun-20 | Lou_39 | 34.5 | 68.0629 | 87.6883 | 25339 |
| Kentucky | S4 | 9-Jun-20 | Lou_43 | 34.6 | 58.5395 | 92.2413 | 21876 |
| Kentucky | S5 | 14-May-20 | Lou_6 | 33.2 | 296.939 | 99.1233 | 110803 |
| Kentucky | S5 | 9-Jun-20 | Lou_38 | 31.4 | 393.77 | 99.0928 | 146800 |
| Kentucky | S6 | 9-Jun-20 | Lou_42 | 33.7 | 57.09 | 92.0856 | 21266 |
| Kentucky | S7 | 23-Apr-20 | Lou_3 | 33.2 | 63.1731 | 84.0764 | 23501 |
| Kentucky | S8 | 21-May-20 | Lou_13 | 34.8 | 148.323 | 98.5546 | 55410 |
| Kentucky | S9 | 23-Apr-20 | Lou_1 | 29.4 | 206.044 | 98.7577 | 76835 |
| Massachusetts | MA1 | 27-May-20 | Jac_89 | 32.8 | 89.2101 | 97.6236 | 33207 |
| New Jersey | NJ1 | 3-May-20 | Jac_04 | 31.2 | 62.1934 | 88.3518 | 23228 |
| New Jersey | NJ1 | 11-May-20 | Jac_30 | 32.6 | 1768.26 | 99.0759 | 658845 |
| New Mexico | NM1 | 6-May-20 | Jac_09 | 30.8 | 14.5232 | 42.8015 | 5435 |
| New Mexico | NM1 | 13-May-20 | Jac_31 | 33 | 127.887 | 98.1686 | 47610 |
| New Mexico | NM1 | 21-May-20 | Jac_69 | 34.3 | 139.456 | 94.8681 | 52042 |
| New Mexico | NM1 | 27-May-20 | Jac_90 | 34.1 | 223.602 | 98.321 | 83229 |
| Oregon | OR1 | 27-May-20 | Jac_92 | 34.7 | 9.50418 | 27.8291 | 3568 |

**Figure 1:**
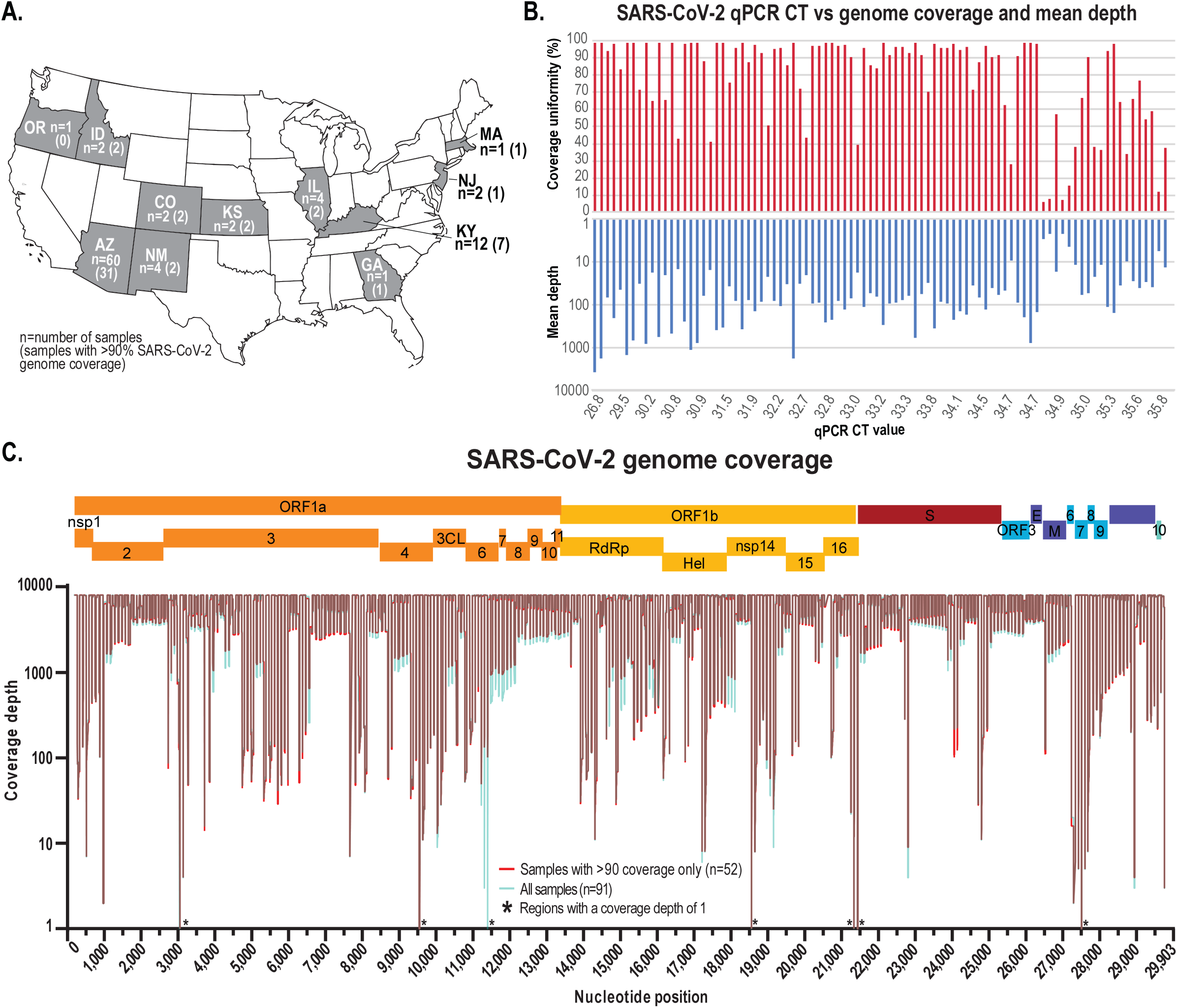
**A**. Map of the United States of America with states where wastewater samples were collected for this study highlighted in grey. **B**. SARS-CoV-2 RT-qPCR Ct detection value for each sample and the corresponding SARS-CoV-2 genome coverage uniformity from the tiling amplicon-based HTS. **C**. SARS-CoV-2 genome coverage of the high-throughput sequencing of all the wastewater samples (cyan) and those with >90% coverage (red). ∗ indicates that these sites have a coverage depth of 1.

### 2.2. Wastewater sample processing and RNA extraction

Aliquots of 150 ml of each composite wastewater sample were filtered through a 0.45 μm polyethersulfone (PES) filter and then subsequently through a 0.2 μm (PES) filter. The filtrate was then concentrated using the Amicon^®^ Ultra 15 Centrifugal Filter Units (MilliporeSigma, USA) by centrifuging at 4500 rpm for 15 min. For each sample, the process was repeated five times in total using two filter units, and subsequently the concentrates were pooled per sample (from the two filter units). For each sample, a 200 μl aliquot was used to extract total RNA using the RNeasy mini kit (Qiagen, USA).

### 2.3. SARS-CoV-2 RT-qPCR detection and high throughput sequencing of SARS-CoV-2 genome sequences

To determine the presence of SARS-CoV-2 in wastewater samples, the extracted RNA was used in a reverse transcription-quantitative PCR (RT-qPCR) assay targeting the E gene, as designed and validated by Corman et al. (2020) and cited by the World Health Organisation (WHO) (WHO, 2020a). This probe-based assay was performed as per the specifications outlined in Corman et al. (2020) using the SuperScript III Platinum One-Step qRT-PCR Kit (Invitrogen, USA). This assay was validated and used by Holland et al. (2020) on SARS-CoV-2 clinical samples.

91 samples from 11 states in the USA (Figure 1) were collected between 7^th^ April 2020 and 16^th^ June 2020 that tested positive, and one negative control sample collected in October 2019 in Tempe, Arizona (Table 1) were selected for sample processing and high-throughput SARS-CoV-2 amplicon sequencing. The SARS-CoV-2 RT-qPCR assay Ct values ranged from 26.8 to 36 for the 91 samples. Total RNA (11 μl) from each sample was used to generate cDNA using the Superscript^®^ IV First-Strand Synthesis System (Thermo Fisher, USA). The manufacturer’s protocol was followed, with one modification, the reverse transcription incubation step (50°C) was increased from 10 to 50 min. 10 μl of cDNA from each sample was used to generate Illumina sequencing libraries (92 libraries in total) with the Swift Nomalase^®^ Amplicon SARS CoV-2 Panel (SNAP) and these were subsequently normalized, pooled and sequenced at Psomagen (USA) on an Illumina HiSeq 2500 sequencer (2×100 paired-end option on 1 lane in rapid mode).

### 2.4. Bioinformatics pipeline and analyses

The Illumina raw read sequences were aligned to the reference genome of SARS-CoV-2 (MN908947; RefSeq ID NC_045512.2) using the Burrows-Wheeler Alignment tool (BWA) MEM (Li and Durbin, 2009). The primers used for the tiling PCR-based amplification step were soft-clipped using iVAR trim tool (Grubaugh et al., 2019) which also removed reads <30nts and reads that started outside of the primer region. Trimming with a sliding window of 4 for a minimum PHRED quality of 20 was performed as default by iVAR. Primers that may have mismatches with the reference sequence were also evaluated and reads from those amplicons with varying primer binding efficiency were also removed as described by Grubaugh et al. (2019). The genome coverage (minimum quality of 20 and 10× coverage) and mean depth was calculated for all samples. Variant calling was performed using iVAR (Grubaugh et al., 2019) with minimum base quality of 20 and 20× coverage with no cut-off frequency since we have population-level sequence data. From the variants that were identified, only those with a p-value <0.05 in the Fisher’s exact test implemented in iVAR (tests if SNV frequency is higher than the mean error rate at the specific position) were maintained. Suggested masked sites due to biases shown by phylogenetic analysis or sequencing technology (De Maio et al., 2020) as of September 2020 were removed for downstream analyses. To identify the novel SNVs, the obtained SNVs from the 52 wastewater samples with SARS-CoV-2 read coverage >90% were searched in the clinical data available in GISAID (Elbe and Buckland-Merrett, 2017; Shu and McCauley, 2017) at two time points (17^th^ June 2020 and 20^th^ November 2020). Variants that were not present in the GISAID deposited SARS-CoV-2 genomes were considered novel, however, to be more stringent, variants that were only present in one of the wastewater samples were removed from further analyses.

### 2.5. Support for lineages assigned by PANGOLIN

Each environmental sample was compared against the SARS-CoV-2 genomes available in GISAID (Elbe and Buckland-Merrett, 2017; Shu and McCauley, 2017), an open-access genomic database, to collect a set of clinical genomes whose mutations were supported by the SNVs identified above. To reduce false positives, basal genomes, defined as those with 3 or fewer mutations relative to the reference (MN908947) were excluded. The set of genomes supported by each environmental sample SNV profile were grouped by lineages assigned by PANGOLIN (Rambaut et al., 2020a) and lineages with fewer than 3 genomes were excluded to avoid any misannotations resulting in false positives. PANGOLIN is an online platform that assigns lineages to sequences (Rambaut et al., 2020a) and is updated as new metadata are submitted to GISAID. For each group of genomes (grouped per PANGOLIN), we then looked to see whether any genome was from North America and, if so, recorded the time between the genome’s sampling date and the collection date of the environmental sample. Note that the set of genomes which we summarize as certain SARS-CoV-2 lineages assigned by PANGOLIN may be different for each environmental sample, and thus the time between clinical and environmental sampling dates depends on the particular SNV profile of the environmental sample. Given that linkage of SNVs is not possible via short read sequencing, support for mutation profiles observed in clinical genomes (and, correspondingly, PANGOLIN) does not guarantee that the lineages were present in the environmental sample.

### 2.6. Sample-based SARS-CoV-2 sequence distance calculation and ordination analysis

The ‘genotype’ of each sample was represented in a four-column matrix. In this matrix, each row corresponds to a position in the reference genome, and the value at each column is the frequency of occurrences for each nucleotide (A, C, G and T). At each genomic position, the Yue & Clayton measure of dissimilarity index (Yue and Clayton, 2005) on the nucleotide frequency of the compared samples was calculated. If the nucleotide frequency at a position of a sample cannot be calculated due to zero depth, the Yue & Clayton measure of dissimilarity index at this position between this sample and any other sample compared is assumed to be zero. The sum of the Yue & Clayton dissimilarity (Yue and Clayton, 2005) of all genomic positions was used as a measure of distance between samples. The distance matrix was constructed by calculating pairwise distances of all samples and was subsequently used for principal coordinates analysis (PCoA) (Gower, 1966).

## 3. Results and discussion

### 3.1. Sample collection, processing and SARS-CoV-2 RT-qPCR screening

Sixty of our 91 samples (66%) were collected in Arizona (9 locations located in Maricopa County, Arizona Sup Figure 1), 12 (13%) were collected from 9 locations in Louisville, Kentucky (Sup Figure 1), and 19 (21%) were collected from other states, see Table 1 and Figure 1A for details.

A sample collected in October 2019 in Tempe, Arizona was processed as a negative control. The samples were processed using a virus concentration approach, followed by RNA extraction and screening for the SARS-CoV-2 by RT-qPCR targeting the E gene. A standard curve with SARS-CoV-2 synthetic RNA (Twist Bioscience, USA) was used to estimate viral load and to establish the limit of detection. Based on the standard curve we determined a consistent limit detection with a Ct-value of ∼34.0. For the samples we analysed, the Ct-values ranged from 26.8 to 36 (Table 1, Figure 1B).

### 3.2. Amplification and high-throughput sequencing of SARS-CoV-2 from wastewater samples

The tiling PCR amplification enrichment process for the SARS-CoV-2 genome generated 341 amplicons covering ∼99% of the genome albeit missing the 200 nts of 5’ end and 162 nts from 3’ end. The genome coverage calculated for all samples ranged between ∼1.3% and ∼99%. 52 of the 91 RT-qPCR positive samples showed >90% coverage (minimum quality of 20 and >10 reads per position) (Table 1). We note that there is no clear correlation between coverage and Ct values obtained using the RT-qPCR assay (Figure 1). This has been shown in other wastewater-derived viral sequencing projects using an Illumina sequencing platforms via an amplification process (Izquierdo Lara et al., 2020) and a capture approach (Crits-Christoph et al., 2021). This lack of correlation is not unexpected given the nature of wastewater, where dilution and degradation play a significant role, thereby this likely results in samples with differing levels of genomic RNA degradation. Furthermore, since the RT-qPCR assay only targets a specific small region of the genome, the Ct-value based quantification vary. Additionally, it is important to highlight that there are variabilities attributed to the handling and transport process of the wastewater samples prior to concentration and RNA extraction.

### 3.3. Wastewater-derived SARS-CoV-2 sequence analyses

For the 52 samples with >90% genome coverage, SNV analysis was undertaken using the program iVAR (minimum quality of 20 and >20 reads per position) without a frequency threshold in order to detect all variations at a population level. This approach was used because, unlike the case with a clinical sample from a single infected individual, wastewater contains material from a population that inhabits a particular region and therefore represents a collection of SARS-CoV-2 variants actively shed by infected individuals within the population. The detected SNVs with *p*-value >0.05 in the Fisher’s exact test were excluded and also *a priori* suggested masked sites due to biases shown by phylogenetic analysis and sequencing technology (De Maio et al., 2020) were excluded from this analysis.

A total of 7973 SNVs were detected for the 52 analysed samples after quality control steps from which the number of detected SNVs per sample ranged from 24 to 793 (Supp. Table 1, Figure 2A). As expected, mean depth is correlated with the number of SNVs detected in each sample (Figure 2B), the regression analysis indicates the trend.

**Figure 2:**
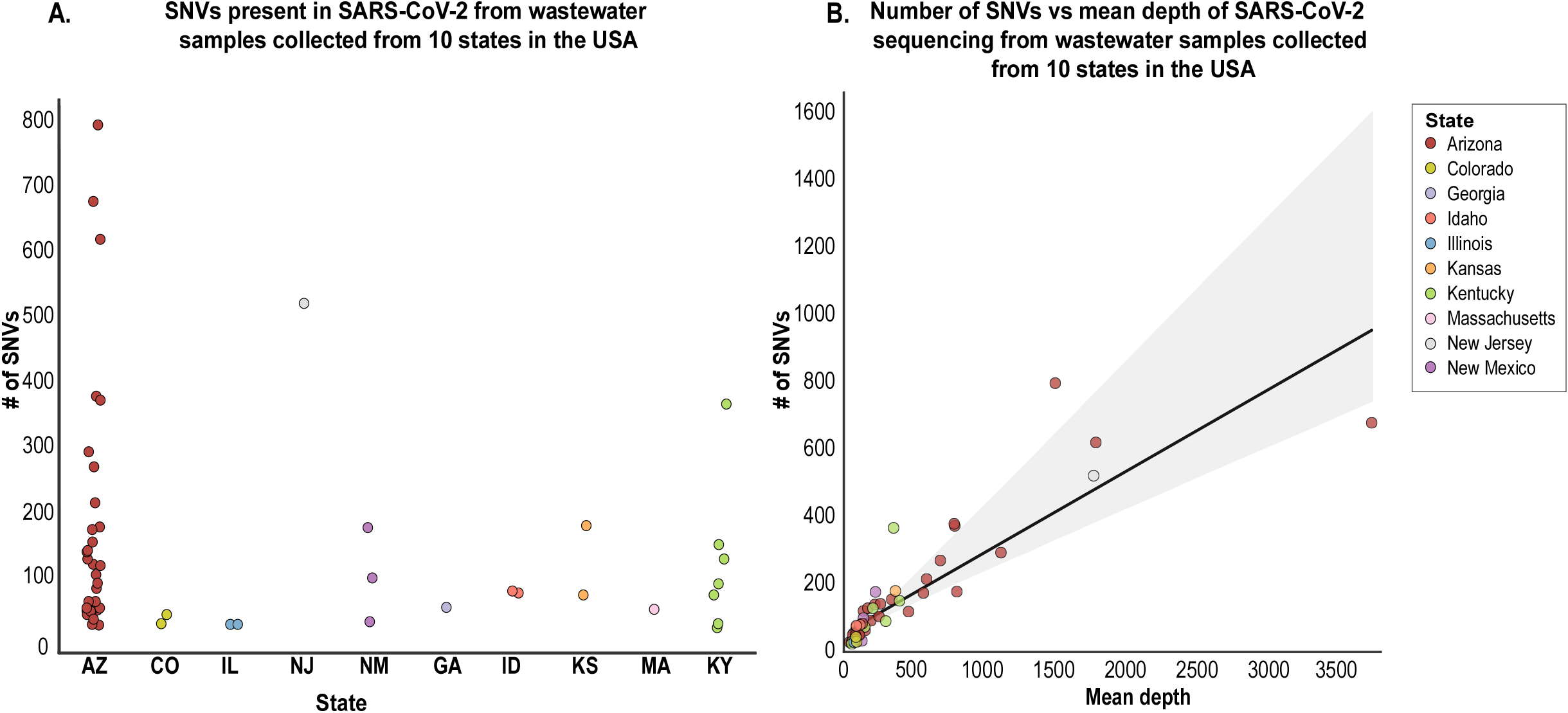
**A**. Number of single nucleotide variants (SNV) per sample across 10 states (each state is represented by a different colour). **B**. Regression analysis, with 95% confidence interval, of the number of wastewater-derived SARS-CoV-2 SNVs detected versus the mean depth for each of the 52 samples with >90% coverage that were analysed. The colour code indicates the states in which the samples were collected.

To determine unique variants within the 52 wastewater-derived SARS-CoV-2 sequences, SNVs counted in more than one sample at each site were removed and this resulted in 5680 unique SNVs identified across the genome. Of these, 4372 are non-synonymous and 1308 are synonymous substitutions. Additionally, 246 are nonsense mutations and 64 are in non-coding regions. We highlight that SNV A23403G responsible for the spike protein substitution D614G that is frequently seen in clinical data, although it has not thus far been shown to be under strong positive selection (Volz et al., 2021), was present in all 52 wastewater-derived SARS-CoV-2 sequences. From one sample (sample #147, Tempe, Arizona), a new variant A23403T was also identified that results in a D614V substitution in the spike protein, but at very low frequency (Sup. Table 1).

### 3.4. Comparative analysis of SARS-CoV-2 SNVs in clinical and wastewater-derived samples during the collection period

The wastewater-derived SARS-CoV-2 SNVs were compared with substitutions that have been detected in clinical-derived sequences. The first aim was to identify possible “novel” SNVs present in the analysed wastewater samples that had not yet been identified in any of the sequences available in GISAID (Elbe and Buckland-Merrett, 2017; Shu and McCauley, 2017) from clinical samples globally. To accomplish this, we initially undertook an analysis to identify all the detected SNVs in the clinical data available from GISAID up until the 17^th^ June 2020 (subsequent to the last day of wastewater sampling in this study - 16^th^ June 2020) which on that date consisted of 45,836 SARS-Cov-2 genome sequences. A total of 548 novel SNVs were identified in the 52 wastewater samples collectively, of these 469 were non-synonymous (not including nonsense mutations) and 79 were synonymous substitutions (Figure 3). Since we evaluated all variants regardless of frequency, some locations (as expected) had more than one possible variant and are illustrated in Figure 3 and outlined in Sup Table 1. These 548 SNVs are distributed along the SARS-CoV-2 genome with three of those located in non-coding regions. The vast majority of “novel” SNVs were detected in up to 8 of the wastewater samples analysed. The exceptions are four non-synonymous mutations, three on the ORF1ab and one in the N gene that are present in >8 samples (Figure 3 and Sup table 1).

**Figure 3:**
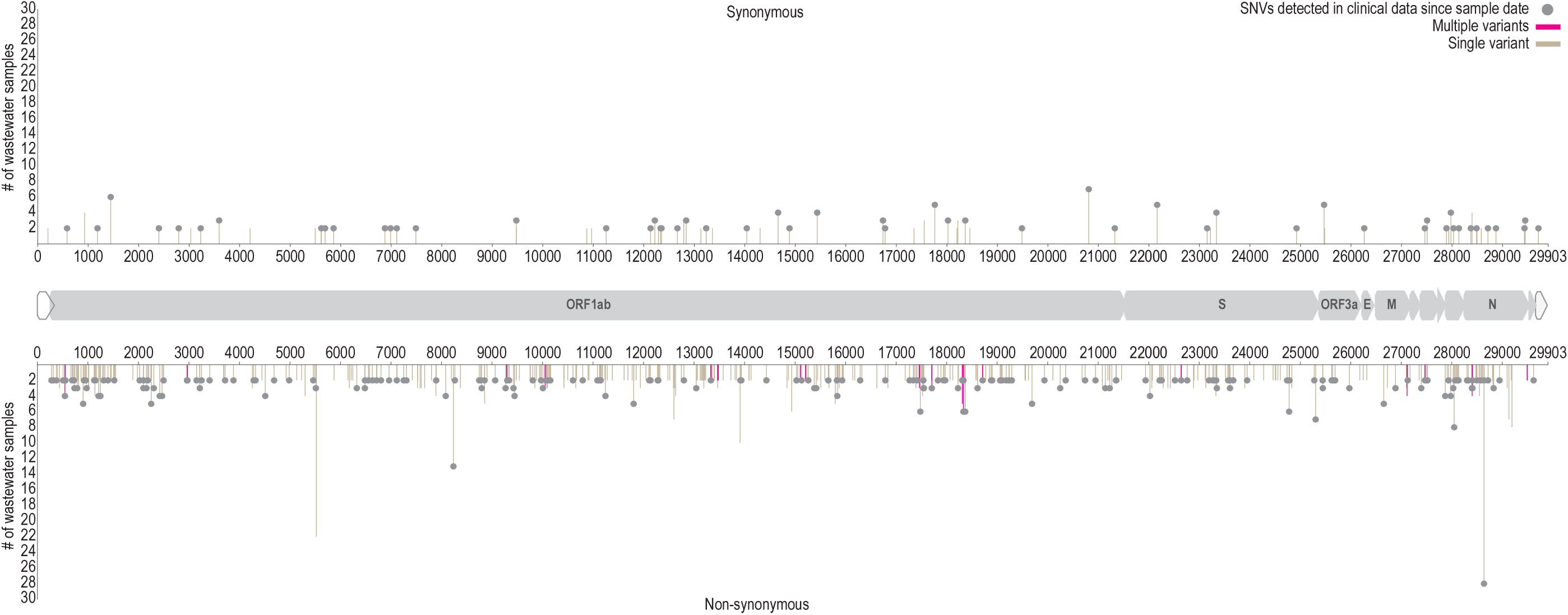
Novel SARS-CoV-2 SNVs (*i*.*e*. not yet detected in clinical-derived samples as of 17th June 2020) identified in the 52 wastewater samples analysed. On the y-axis are the number of samples containing the SNV and on the x-axis is the relative position of SNV in the SARS-CoV-2 genome. Positions with multiple variants are marked in red and those marked with grey circles represent the SNVs that have been detected up until 20th November 2020 in clinical samples.

### 3.5. Identification of SARS-CoV-2 SNVs in wastewater samples in clinical-derived samples post-collection period

To determine how many SNVs have been identified post wastewater sample collection (16^th^ June 2020), a second SNV comparison was performed with all the clinical-derived sequence data available as of 20^th^ November 2020 (203,741 SARS-Cov-2 genomes available at GISAID). Based on the analysis of samples during the collection period, SNVs that were not detected in the clinical-derived sequence data were considered as novel SNVs. From the 548 SNVs considered as “novel” from the wastewater-derived samples, 263 SNVs have subsequently been identified in clinical-derived samples in the period of 17^th^ June - 20^th^ November 2020 (Sup Table 1, Figure 3). 285 SNVs identified in the wastewater-derived samples with the last sampling date of 16^th^ June 2020 have not been identified in clinical-derived SARS-CoV-2 sequences between then and 20^th^ November 2020.

It is important to highlight that the detection of these “novel” SNVs does not necessarily indicate they are fixed in SARS-CoV-2 lineages that are actively being transmitted nor is it possible to determine if any of these SNVs are linked within lineages. Nonetheless, the identification of the “novel” SNVs clearly demonstrates the relevance of wastewater-derived SARS-CoV-2 sequence analysis which can provide valuable information on SNVs that are not captured using clinical-derived approaches. The wastewater-derived sequence analysis does provide information at a population scale and can allow for rapid detection of clinically relevant / important SNVs.

### 3.6. Determination of putative lineages of SARS-CoV-2 in wastewater-derived sequences

Given that wastewater harbours a collective population of SARS-CoV-2 and therefore likely many variants, it is not ideal to determine consensus sequences and consensus sequences-based phylogeny. Therefore, our first approach was to evaluate which clades in the global phylogeny of clinical-derived sequences are supported by the SNVs present in each sample based on the SARS-CoV-2 lineages assigned by PANGOLIN (Rambaut et al., 2020a). The represented SARS-CoV-2 lineages for each wastewater sample that are supported are shown in Figure 4. We determined the time frames for which these lineages were first detected in North American clinical-derived sequences relative to the date each wastewater sample was collected (Figure 4A).

**Figure 4:**
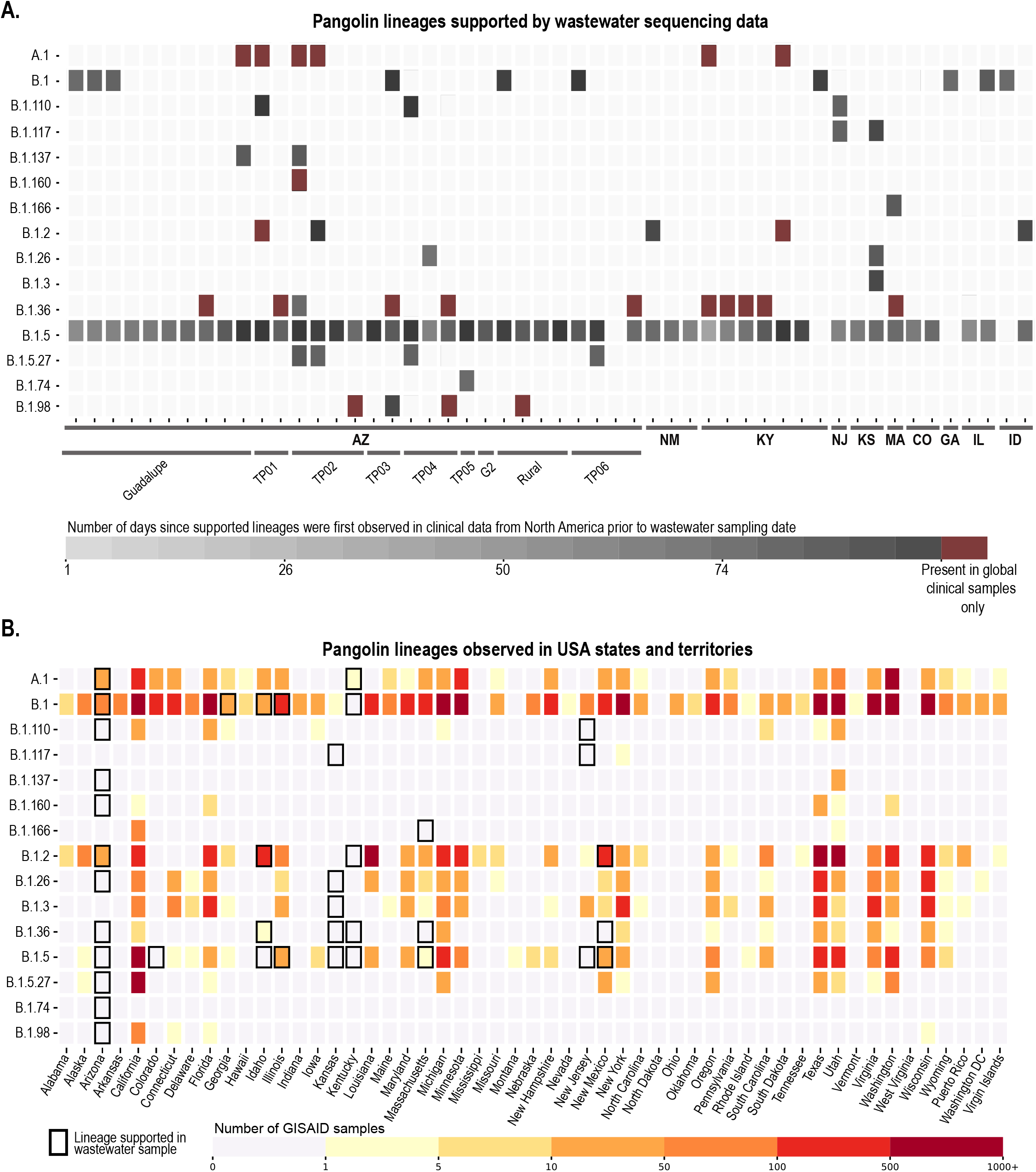
Publicly available genomes from clinically derived data deposited in GISAID, grouped by PANGOLIN, whose mutations were consistent with those observed in wastewater samples. **A**. Heatmap showing the number of days between sample collection and when supported lineages were first observed in clinical data. Each wastewater sample (52 samples across 10 states) contained support for different clinical samples which are grouped here by PANGOLIN, some of which have only been observed outside North America (indicated as “global only”). **B**. Clinical genomes reported in USA states and territories which were assigned to PANGOLIN supported by at least one environmental sample. Black borders indicate lineages supported in environmental samples from the respective location.

We also undertook a comprehensive analysis of all the lineages detected in each state in the USA up to November 2020 that were supported by at least one environmental sample, this included the number of clinical-derived SARS-CoV-2 genomes sequenced in each lineage (Figure 4B). This approach helps to determine whether wastewater-based surveillance for SARS-CoV-2 can provide valuable insights on putative circulating lineages in the wastewater contributing population. Although there are several limitations to the analysis of wastewater-derived SARS-CoV-2 sequences, our analysis of SNV-based supported lineages revealed some interesting findings. From the 52 analysed wastewater samples, 15 SARS-CoV-2 lineages assigned by PANGOLIN (Rambaut et al., 2020a) were supported, with lineage B.1.5 being the most prominent for the wastewater-derived sequences. The B.1.5 lineage has been identified in clinical samples in 27 USA states. Our wastewater-derived sequence data suggests that B.1.5 may also be present in 6 additional states in the USA (Arizona, Colorado, Idaho, Kansas, Kentucky and New Jersey). In 17 of the 52 wastewater samples, there were up to two supported SARS-CoV-2 lineages that had not been detected in North American clinical samples, during the period of our wastewater collection, as of 17^th^ June 2020 (Figure 4). These 17 samples were from the states of Arizona, Kentucky and Massachusetts (Figure 4B). In wastewater-derived sequences from Arizona, which represents the greatest proportion of samples, the observed circulating lineages based on clinical-derived sequences are well represented (Ladner et al., 2020), with an additional nine possible circulating lineages identified.

Although wastewater-based SARS-CoV-2 sequence analysis does not provide the same level of genome confidence (and thus lineage assignment) as those from clinical samples, the wastewater-derived data can be used to identify possible circulating lineages and assess the diversity of SARS-CoV-2. We would like to emphasize that despite us identifying supported lineages based on SNVs analysis, without verification of full genomes using long read sequencing technologies it is not possible to confirm all the specific lineages present in the wastewater. Nevertheless, it is apparent that valuable population-level variant information on SARS-CoV-2 can be gleaned from wastewater sampling, including significant sequence data that are potentially missed in clinical-derived sequence data where genomes are sequenced from predominantly infected individuals who might represent a small percentage of those shedding virus in a community.

### 3.7. Principal coordinates analysis (PCoA) analysis of nucleotide frequencies to diversity estimate

In Figure 5, we show our PCoA analysis results using nucleotide frequencies to evaluate the viral population diversity within and between samples. SARS-CoV-2 sequences in the samples from the ten states were overall highly diverse, and those with two or more samples from the same state tend to cluster closer together (Figure 5). The main exceptions are those from Kansas (20^th^ May 2020 and 27^th^ May 2020) and Colorado (20^th^ May 2020 and 28^th^ May 2020) that do not cluster together, both were collected a week apart, and the locations have an estimated human population size of ∼25,900 and ∼8,300, respectively. Additionally, the Arizona wastewater SARS-CoV-2 sequences are broadly distributed in the PCoA plot which is likely a consequence of the large number of samples collected over a three-month period across several sites within Maricopa County, Arizona (Tempe sites, Guadalupe and Gilbert) (Figure 5A, B and C). In comparison to those in the Arizona wastewater samples, the SARS-CoV-2 sequences in samples from Louisville (Kentucky) are much more tightly clustered in the PCoA plot despite sampling from several locations in the city over a two-month period (Figure 5A). Despite the large number of samples collected in Arizona compared to Kentucky, and the other states, if seven individual samples were to be randomly picked from each location over the same period as those from Kentucky the SARS-CoV-2 genetic distance between them would still be apparently higher for Arizona. We hypothesize that one contributing factor to the differences in viral diversity present in these two areas *i*.*e*. Maricopa County Arizona and Louisville (Kentucky), is that, Tempe (the region where the majority of the samples were collected) is home to one of the largest universities in the USA, Maricopa County is the 4^th^ most populous county in the USA with ∼4.4 million inhabitants (Maricopa County 2020) and a major travel hub with an international airport.

**Figure 5:**
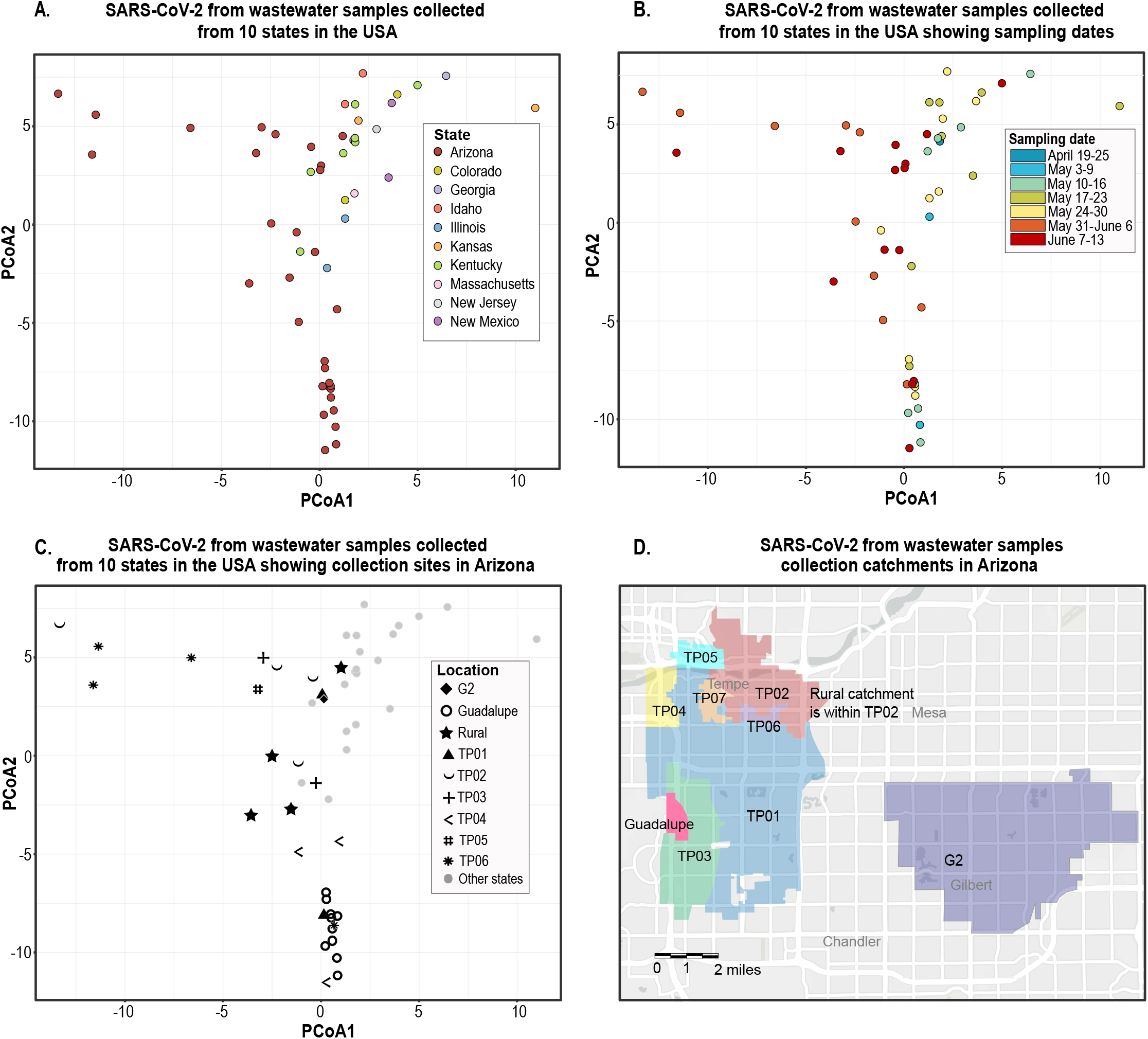
Principal coordinate analysis (PCoA) of SARS-CoV-2 sequence data derived from wastewater samples. **A**. Distribution of sequences from samples collected in ten states (each represented by a different colour) in the USA showing pairwise distance based on genomic composition between viral populations present in each sample. **B**. Timeline representation (shown by the colour gradient) of samples taken from the sample locations across ten USA states between April-June 2020 with pairwise distance based on genomic composition between viral populations present in each sample. **C**. Spatial representation of SARS-CoV-2 sequences from samples collected from various regions within Arizona (represented by different symbols) comparative to those from other states. **D**. Sampling catchments in Tempe, Guadalupe and Gilbert, Arizona.

The highest number of samples collected within a state both temporally and spatially for this study was in Arizona. In Arizona, we note that the wastewater-derived SARS-CoV-2 sequences in samples from the same locations do not necessarily cluster together in the PCoA plot (Figure 5C). Nonetheless, there are clear shifts in the SARS-CoV-2 sequence variants in each location over time (Figure 5B and C). This is most evident for the Town of Guadalupe (Arizona) given the sampling effort here, where the SARS-CoV-2 sequences in the samples collected in early May 2020 cluster with lower distance but we can see a clear shift in the viral population starting late May 2020 through to early June (Figures 5B and C) which coincides with stay at home lockdown being lifted on 15^th^ May 2020. It is important to highlight that the Town of Guadalupe (Arizona) has a small resident community (∼6,500) from where wastewater was collected. Moreover, SARS-CoV-2 sequences in the samples from the same location at closer timepoints are often more likely to be similar, yet there are exceptions such as the samples from site TP04 (Tempe, Arizona) that have no resident population (Figure 5B and C). The shift in SARS-CoV-2 sequence diversity in locations such as TP04 (Tempe, Arizona) over time may be due to new infections given the transient population.

Increases in SARS-CoV-2 viral RNA in wastewater have been correlated to an increase in the number of cases locally (Medema et al., 2020). Observing a shift in the SARS-CoV-2 population diversity through wastewater analysis with time provides insights into corresponding dynamics of increased infection in the community. For example, in Tempe, the number of recorded cases nearly doubled in June 2020. When analysing wastewater-derived SARS-CoV-2 sequence data and correlating it with human dynamics, business districts in the cities will certainly see the activity of transient community members and this will likely reflect in sequence diversity data.

## 4. Conclusion

Wastewater-based analysis is rapidly becoming a useful platform for investigating the epidemiology of viruses shed in human excretions (Farkas et al., 2018; Farkas et al., 2020; Tambini et al., 1993). In this study, we analyse HTS data of wastewater-derived SARS-CoV-2 sequences to determine SNVs, putative circulating lineages and also population structure at a spatial and temporal scale. Analysis of wastewater-derived SARS-CoV-2 sequences from 10 states (Figure 2A) highlighted that the SNVs range from 24 to 793 SNVs for each sample with the highest number in samples from Arizona. As expected, mean depth is correlated with the number of SNVs detected in each sample (Figure 2B). Our major findings included the detection of a high number of novel SNVs detected (548) in the 52 wastewater-derived SARS-CoV-2 sequences analysed here (Figure 3) that had not been identified in clinical samples previously to the last day of our sampling (16^th^ June 2020). Furthermore, 263 SARS-CoV-2 SNVs identified in wastewater samples sampled during our collection period had not been identified in clinical-derived sequences as of 20^th^ November 2020 (Figure 3). It is likely that a large proportion of these SNVs are in “actively circulating” viruses and could have some biological significance.

Through analysis of SNVs in the SARS-CoV-2 sequences in each wastewater sample, we were able to identify putative Phylogenetic Assignment of Named Global Outbreak Lineages (PANGOLIN) that are known to be circulating in the USA as well as several lineages that had not been detected in North America up until 20^th^ November 2020. For the samples from the states of Arizona and Kentucky where we had undertaken temporal and spatial sampling, some PANGOLIN that had been detected in SARS-CoV-2 clinical-sequence data were also supported in the wastewater in addition to several other putative lineages which may have been missed by clinical sampling (Figure 4). In conjunction with diversity analyses using distance matrices (Figure 5) this shows trends in viral populations which can help to track the spread of the SARS-CoV-2.

This study supports the use of wastewater sampling as a tool suitable for analysing the genomics of ongoing outbreaks of infectious diseases, such as SARS-CoV-2. As demonstrated here, HTS of RNA from wastewater can provide novel information on SNVs and lineages which, when coupled with that derived from clinical data, can help identify new emerging variants/lineages of clinical importance within a population. The study results indicating a shift in the SARS-CoV-2 sequence variation in wastewater from each location over time shows the ongoing need for such approaches. As a collective, the approaches we have outlined in this study can be used within a public health setting as an early warning tool to inform infectious disease mitigation measures.

## Sequence data

Sequences are deposited in NCBI’s SRA under the project number PRJNA662596; SRA # SRR12618464 - SRR12618554 and SRR13289969.

## Data Availability

Sequence data
Sequences are deposited in NCBI SRA under the project number PRJNA662596; SRA # SRR12618464 - SRR12618554 and SRR13289969.

## Acknowledgements

The research reported in this publication was supported by the National Library of Medicine of the National Institutes of Health under Award Number U01LM013129 to RUH, MS and AV. The work of XJ was supported by the Intramural Research Program of the National Library of Medicine, National Institutes of Health. The content is solely the responsibility of the authors and does not necessarily represent the official views of the National Institutes of Health. The work in Louisville, KY, was supported in part by grants from the James Graham Brown Foundation and the Owsley Brown Family Foundation. The authors would like to thank William Mancini (Enterprise GIS & Data Analyst, City of Tempe, USA) for providing the base map for Tempe.

## Conflicts of Interest

E.M.D and R.U.H. are cofounders of AquaVitas, LLC, 9260 E. Raintree, Ste 130, Scottsdale, AZ 85260, USA, an ASU start-up company providing commercial services in wastewater-based epidemiology. R.U.H. is the founder of OneWaterOneHealth, a non-profit project of the Arizona State University Foundation.

## Figure legends and table text

**Supplementary Figure 1:** Wastewater sampling catchments in Louisville (Kentucky), Sites 1 and 7 represent collection sites of hospitals and Site 9 is a sewer district facility.

**Supplementary Table1**. Summary of the SNVs detected in SARS-CoV-2 sequences in the 52 wastewater samples (*n*=7,973). In the order of the table, the information contained in each column is: the sample name, date of collection, state, location within the state, SNV position, reference nucleotide, alternative nucleotide, frequency of alternative nucleotide, total read depth at position, reference codon, reference amino acid, alternative codon, alternative amino acid, bin (number of wastewater samples that contain that SNV), global frequency of SNV, USA frequency of SNV and if the SNV is synonymous (syn) or non-synonymous (Nsyn).

## Notes

### Competing Interest Statement

The authors have declared no competing interest.

### Author Declarations

Arizona State University Institutional Review Board

## References

1. Adams, E.R., Ainsworth, M., Anand, R., Andersson, M.I., Auckland, K., Baillie, J.K., Barnes, E., Beer, S., Bell, J.I. and Berry, T. 2020. Antibody testing for COVID-19: A report from the National COVID Scientific Advisory Panel. Wellcome Open Research 5(139), 139.https://doi.org/10.12688/wellcomeopenres.15927.1.

2. Ahmed, W., Angel, N., Edson, J., Bibby, K., Bivins, A., O’Brien, J.W., Choi, P.M., Kitajima, M., Simpson, S.L., Li, J., Tscharke, B., Verhagen, R., Smith, W.J.M., Zaugg, J., Dierens, L., Hugenholtz, P., Thomas, K.V. and Mueller, J.F. 2020. First confirmed detection of SARS-CoV-2 in untreated wastewater in Australia: A proof of concept for the wastewater surveillance of COVID-19 in the community. Sci Total Environ 728, 138764.https://doi.org/10.1016/j.scitotenv.2020.138764.

3. Andersen, K.G., Rambaut, A., Lipkin, W.I., Holmes, E.C. and Garry, R.F. 2020. The proximal origin of SARS-CoV-2. Nat Med 26(4), 450–452.https://doi.org/10.1038/s41591-020-0820-9.

4. Balboa, S., Mauricio-Iglesias, M., Rodriguez, S., Martínez-Lamas, L., Vasallo, F.J., Regueiro, B. and Lema, J.M. 2020. The fate of SARS-CoV-2 in wastewater treatment plants points out the sludge line as a suitable spot for incidence monitoring. medRxiv 10.1101/2020.05.25.20112706.https://doi.org/10.1101/2020.05.25.20112706.

5. Becker, M., Strengert, M., Junker, D., Kerrinnes, T., Kaiser, P.D., Traenkle, B., Dinter, H., Haering, J., Zeck, A., Weise, F., Peter, A., Hoerber, S., Fink, S., Ruoff, F., Bakchoul, T., Baillot, A., Lohse, S., Cornberg, M., Illig, T., Gottlieb, J., Smola, S., Karch, A., Berger, K., Rammensee, H.-G., Schenke-Layland, K., Nelde, A., Maerklin, M., Heitmann, J.S., Walz, J.S., Templin, M.F., Joos, T.O., Rothbauer, U., Krause, G. and Schneiderhan-Marra, N. 2020. Going beyond clinical routine in SARS-CoV-2 antibody testing - A multiplex corona virus antibody test for the evaluation of cross-reactivity to endemic coronavirus antigens. medRxiv 2020.07.17.20156000.https://doi.org/2020.07.17.20156000.

6. Boni, M.F., Lemey, P., Jiang, X., Lam, T.T., Perry, B.W., Castoe, T.A., Rambaut, A. and Robertson, D.L. 2020. Evolutionary origins of the SARS-CoV-2 sarbecovirus lineage responsible for the COVID-19 pandemic. Nat Microbiol 5(11), 1408–1417.https://doi.org/10.1038/s41564-020-0771-4.

7. Bryant, J.E., Azman, A.S., Ferrari, M.J., Arnold, B.F., Boni, M.F., Boum, Y., Hayford, K., Luquero, F.J., Mina, M.J., Rodriguez-Barraquer, I., Wu, J.T., Wade, D., Vernet, G. and Leung, D.T. 2020. Serology for SARS-CoV-2: Apprehensions, opportunities, and the path forward. Sci Immunol 5(47).https://doi.org/10.1126/sciimmunol.abc6347.

8. Buitrago-Garcia, D., Egli-Gany, D., Counotte, M.J., Hossmann, S., Imeri, H., Ipekci, A.M., Salanti, G. and Low, N. 2020. Occurrence and transmission potential of asymptomatic and presymptomatic SARS-CoV-2 infections: A living systematic review and meta-analysis. PLoS Med 17(9), e1003346.https://doi.org/10.1371/journal.pmed.1003346.

9. Byambasuren, O., Cardona, M., Bell, K., Clark, J., McLaws, M.-L. and Glasziou, P. 2020. Estimating the extent of asymptomatic COVID-19 and its potential for community transmission: systematic review and meta-analysis. medRixv 10.1101/2020.05.10.20097543.https://doi.org/10.1101/2020.05.10.20097543.

10. CDC 2020a Centers for Disease Control and Prevention - CDC Diagnostic Tests for COVID-19. https://www.cdc.gov/coronavirus/2019-ncov/lab/testing.html

11. CDC 2020b Centers for Disease Control and Prevention - Serology Testing for COVID-19 at CDC. https://www.cdc.gov/coronavirus/2019-ncov/lab/serology-testing.html

12. Chen, Y., Chen, L., Deng, Q., Zhang, G., Wu, K., Ni, L., Yang, Y., Liu, B., Wang, W., Wei, C., Yang, J., Ye, G. and Cheng, Z. 2020. The presence of SARS-CoV-2 RNA in the feces of COVID-19 patients. J Med Virol 92(7), 833–840.https://doi.org/10.1002/jmv.25825.

13. Corman, V.M., Landt, O., Kaiser, M., Molenkamp, R., Meijer, A., Chu, D.K., Bleicker, T., Brunink, S., Schneider, J., Schmidt, M.L., Mulders, D.G., Haagmans, B.L., van der Veer, B., van den Brink, S., Wijsman, L., Goderski, G., Romette, J.L., Ellis, J., Zambon, M., Peiris, M., Goossens, H., Reusken, C., Koopmans, M.P. and Drosten, C. 2020. Detection of 2019 novel coronavirus (2019-nCoV) by real-time RT-PCR. Euro Surveill 25(3).https://doi.org/10.2807/1560-7917.ES.2020.25.3.2000045.

14. Crits-Christoph, A., Kantor, R.S., Olm, M.R., Whitney, O.N., Al-Shayeb, B., Lou, Y.C., Flamholz, A., Kennedy, L.C., Greenwald, H., Hinkle, A., Hetzel, J., Spitzer, S., Koble, J., Tan, A., Hyde, F., Schroth, G., Kuersten, S., Banfield, J.F. and Nelson, K.L. 2021. Genome Sequencing of Sewage Detects Regionally Prevalent SARS-CoV-2 Variants. mBio 12(1).https://doi.org/10.1128/mBio.02703-20.

15. D’Aoust, P.M., Mercier, E., Montpetit, D., Jia, J.J., Alexandrov, I., Neault, N., Baig, A.T., Mayne, J., Zhang, X., Alain, T., Langlois, M.A., Servos, M.R., MacKenzie, M., Figeys, D., MacKenzie, A.E., Graber, T.E. and Delatolla, R. 2021. Quantitative analysis of SARS- CoV-2 RNA from wastewater solids in communities with low COVID-19 incidence and prevalence. Water Res 188, 116560.https://doi.org/10.1016/j.watres.2020.116560.

16. De Maio, N., Walker, C., Borges, R., Weilguny, L., Slodkowicz, G. and Goldman, N. 2020. Issues with SARS-CoV-2 sequencing data. https://virological.org/t/issues-with-sars-cov-2-sequencing-data/473/1

17. Dong, E., Du, H. and Gardner, L. 2020. An interactive web-based dashboard to track COVID- 19 in real time. Lancet Infect Dis 20(5), 533–534.https://doi.org/10.1016/S1473-3099(20)30120-1.

18. Elbe, S. and Buckland-Merrett, G. 2017. Data, disease and diplomacy: GISAID’s innovative contribution to global health. Glob Chall 1(1), 33–46.https://doi.org/10.1002/gch2.1018.

19. Farkas, K., Cooper, D.M., McDonald, J.E., Malham, S.K., de Rougemont, A. and Jones, D.L. 2018. Seasonal and spatial dynamics of enteric viruses in wastewater and in riverine and estuarine receiving waters. Sci Total Environ 634, 1174–1183.https://doi.org/10.1016/j.scitotenv.2018.04.038.

20. Farkas, K., Hillary, L.S., Malham, S.K., McDonald, J.E. and Jones, D.L. 2020. Wastewater and public health: the potential of wastewater surveillance for monitoring COVID-19. Curr Opin Environ Sci Health 17, 14–20.https://doi.org/10.1016/j.coesh.2020.06.001.

21. Gorbalenya, A.E., Baker, S.C., Baric, R.S., de Groot, R.J., Drosten, C., Gulyaeva, A.A., Haagmans, B.L., Lauber, C., Leontovich, A.M., Neuman, B.W., Penzar, D., Perlman, S., Poon, L.L.M., Samborskiy, D.V., Sidorov, I.A., Sola, I., Ziebuhr, J. and Coronaviridae Study Group of the International Committee on Taxonomy of, V. 2020. The species Severe acute respiratory syndrome-related coronavirus: classifying 2019-nCoV and naming it SARS-CoV-2. Nature Microbiology 5(4), 536–544.https://doi.org/10.1038/s41564-020-0695-z.

22. Gower, J.C. 1966. Some Distance Properties of Latent Root and Vector Methods Used in Multivariate Analysis. Biometrika 53(3/4), 325.https://doi.org/10.2307/2333639.

23. Grubaugh, N.D., Gangavarapu, K., Quick, J., Matteson, N.L., De Jesus, J.G., Main, B.J., Tan, A.L., Paul, L.M., Brackney, D.E., Grewal, S., Gurfield, N., Van Rompay, K.K.A., Isern, S., Michael, S.F., Coffey, L.L., Loman, N.J. and Andersen, K.G. 2019. An amplicon-based sequencing framework for accurately measuring intrahost virus diversity using PrimalSeq and iVar. Genome Biol 20(1), 8.https://doi.org/10.1186/s13059-018-1618-7.

24. Holland, L.A., Kaelin, E.A., Maqsood, R., Estifanos, B., Wu, L.I., Varsani, A., Halden, R.U., Hogue, B.G., Scotch, M. and Lim, E.S. 2020. An 81-Nucleotide Deletion in SARS-CoV- 2 ORF7a Identified from Sentinel Surveillance in Arizona (January to March 2020). J Virol 94(14).https://doi.org/10.1128/JVI.00711-20.

25. Izquierdo Lara, R.W., Elsinga, G., Heijnen, L., Oude Munnink, B.B., Schapendonk, C.M.E., Nieuwenhuijse, D., Kon, M., Lu, L., Aarestrup, F.M., Lycett, S., Medema, G., Koopmans, M.P.G. and de Graaf, M. 2020. Monitoring SARS-CoV-2 circulation and diversity through community wastewater sequencing. medRxiv 10.1101/2020.09.21.20198838.https://doi.org/10.1101/2020.09.21.20198838.

26. Jones, D.L., Baluja, M.Q., Graham, D.W., Corbishley, A., McDonald, J.E., Malham, S.K., Hillary, L.S., Connor, T.R., Gaze, W.H., Moura, I.B., Wilcox, M.H. and Farkas, K. 2020. Shedding of SARS-CoV-2 in feces and urine and its potential role in person-to-person transmission and the environment-based spread of COVID-19. Sci Total Environ 749, 141364.https://doi.org/10.1016/j.scitotenv.2020.141364.

27. Kimball, A., Hatfield, K.M., Arons, M., James, A., Taylor, J., Spicer, K., Bardossy, A.C., Oakley, L.P., Tanwar, S., Chisty, Z., Bell, J.M., Methner, M., Harney, J., Jacobs, J.R., Carlson, C.M., McLaughlin, H.P., Stone, N., Clark, S., Brostrom-Smith, C., Page, L.C., Kay, M., Lewis, J., Russell, D., Hiatt, B., Gant, J., Duchin, J.S., Clark, T.A., Honein, M.A., Reddy, S.C., Jernigan, J.A., Public Health, S., King, C. and Team, C.C.-I. 2020. Asymptomatic and Presymptomatic SARS-CoV-2 Infections in Residents of a Long-Term Care Skilled Nursing Facility - King County, Washington, March 2020. MMWR Morb Mortal Wkly Rep 69(13), 377–381.https://doi.org/10.15585/mmwr.mm6913e1.

28. Kocamemi, B.A., Kurt, H., Sait, A., Sarac, F., Saatci, A.M. and Pakdemirli, B. 2020. SARS- CoV-2 Detection in Istanbul Wastewater Treatment Plant Sludges. medRxiv 10.1101/2020.05.12.20099358.https://doi.org/10.1101/2020.05.12.20099358.

29. Kumar, M., Patel, A.K., Shah, A.V., Raval, J., Rajpara, N., Joshi, M. and Joshi, C.G. 2020. First proof of the capability of wastewater surveillance for COVID-19 in India through detection of genetic material of SARS-CoV-2. Sci Total Environ 746, 141326.https://doi.org/10.1016/j.scitotenv.2020.141326.

30. La Rosa, G., Iaconelli, M., Mancini, P., Bonanno Ferraro, G., Veneri, C., Bonadonna, L., Lucentini, L. and Suffredini, E. 2020. First detection of SARS-CoV-2 in untreated wastewaters in Italy. Sci Total Environ 736, 139652.https://doi.org/10.1016/j.scitotenv.2020.139652.

31. Ladner, J.T., Larsen, B.B., Bowers, J.R., Hepp, C.M., Bolyen, E., Folkerts, M., Sheridan, K., Pfeiffer, A., Yaglom, H., Lemmer, D., Sahl, J.W., Kaelin, E.A., Maqsood, R., Bokulich, N.A., Quirk, G., Watts, T.D., Komatsu, K.K., Waddell, V., Lim, E.S., Caporaso, J.G., Engelthaler, D.M., Worobey, M. and Keim, P. 2020. An Early Pandemic Analysis of SARS-CoV-2 Population Structure and Dynamics in Arizona. mBio 11(5).https://doi.org/10.1128/mBio.02107-20.

32. Li, H. and Durbin, R. 2009. Fast and accurate short read alignment with Burrows-Wheeler transform. Bioinformatics 25(14), 1754–1760.https://doi.org/10.1093/bioinformatics/btp324.

33. Maricopa County. 2020 Maricopa County, AZ. https://www.maricopa.gov/

34. Medema, G., Heijnen, L., Elsinga, G., Italiaander, R. and Brouwer, A. 2020. Presence of SARS-Coronavirus-2 RNA in Sewage and Correlation with Reported COVID-19 Prevalence in the Early Stage of the Epidemic in The Netherlands. Environmental Science & Technology Letters 7(7), 511–516.https://doi.org/10.1021/acs.estlett.0c00357.

35. Nemudryi, A., Nemudraia, A., Wiegand, T., Surya, K., Buyukyoruk, M., Vanderwood, K.K., Wilkinson, R. and Wiedenheft, B. 2020. Temporal detection and phylogenetic assessment of SARS-CoV-2 in municipal wastewater. Cell Rep Med 22(16), 1000098.https://doi.org/10.1101/2020.04.15.20066746.

36. Park, S.K., Lee, C.W., Park, D.I., Woo, H.Y., Cheong, H.S., Shin, H.C., Ahn, K., Kwon, M.J. and Joo, E.J. 2020. Detection of SARS-CoV-2 in Fecal Samples From Patients With Asymptomatic and Mild COVID-19 in Korea. Clin Gastroenterol Hepatol 10.1016/j.cgh.2020.06.005.https://doi.org/10.1016/j.cgh.2020.06.005.

37. Peccia, J., Zulli, A., Brackney, D.E., Grubaugh, N.D., Kaplan, E.H., Casanovas-Massana, A., Ko, A.I., Malik, A.A., Wang, D., Wang, M., Warren, J.L., Weinberger, D.M., Arnold, W. and Omer, S.B. 2020. Measurement of SARS-CoV-2 RNA in wastewater tracks community infection dynamics. Nat Biotechnol 38(10), 1164–1167.https://doi.org/10.1038/s41587-020-0684-z.

38. Rambaut, A., Holmes, E.C., O’Toole, A., Hill, V., McCrone, J.T., Ruis, C., du Plessis, L. and Pybus, O.G. 2020a. A dynamic nomenclature proposal for SARS-CoV-2 lineages to assist genomic epidemiology. Nat Microbiol 5(11), 1403–1407.https://doi.org/10.1038/s41564-020-0770-5.

39. Rambaut, A., Loman, N., Pybus, O., Barclay, W., Barrett, J., Carabelli, A., Connor, T., Peacock, T., Robertson, D.L. and Volz, E. 2020b Preliminary genomic characterisation of an emergent SARS-CoV-2 lineage in the UK defined by a novel set of spike mutations.

40. Randazzo, W., Truchado, P., Cuevas-Ferrando, E., Simon, P., Allende, A. and Sanchez, G. 2020. SARS-CoV-2 RNA in wastewater anticipated COVID-19 occurrence in a low prevalence area. Water Res 181, 115942.https://doi.org/10.1016/j.watres.2020.115942.

41. Shu, Y. and McCauley, J. 2017. GISAID: Global initiative on sharing all influenza data - from vision to reality. Euro Surveill 22(13).https://doi.org/10.2807/1560-7917.ES.2017.22.13.30494.

42. Syangtan, G., Bista, S., Dawadi, P., Rayamajhee, B., Shrestha, L.B., Tuladhar, R. and Joshi, D.R. 2020. Asymptomatic people with SARS-CoV-2 as unseen carriers of COVID-19: A systematic review and meta-analysis. Reseach Square 10.21203/rs.3.rs-39512/v1.https://doi.org/10.21203/rs.3.rs-39512/v1.

43. Tambini, G., Andrus, J.K., Marques, E., Boshell, J., Pallansch, M., de Quadros, C.A. and Kew, O. 1993. Direct detection of wild poliovirus circulation by stool surveys of healthy children and analysis of community wastewater. J Infect Dis 168(6), 1510–1514.https://doi.org/10.1093/infdis/168.6.1510.

44. Tang, A., Tong, Z.D., Wang, H.L., Dai, Y.X., Li, K.F., Liu, J.N., Wu, W.J., Yuan, C., Yu, M.L., Li, P. and Yan, J.B. 2020. Detection of Novel Coronavirus by RT-PCR in Stool Specimen from Asymptomatic Child, China. Emerg Infect Dis 26(6), 1337–1339.https://doi.org/10.3201/eid2606.200301.

45. Tegally, H., Wilkinson, E., Giovanetti, M., Iranzadeh, A., Fonseca, V., Giandhari, J., Doolabh, D., Pillay, S., San, E.J., Msomi, N., Mlisana, K., von Gottberg, A., Walaza, S., Allam, M., Ismail, A., Mohale, T., Glass, A.J., Engelbrecht, S., Van Zyl, G., Preiser, W., Petruccione, F., Sigal, A., Hardie, D., Marais, G., Hsiao, M., Korsman, S., Davies, M.-A., Tyers, L., Mudau, I., York, D., Maslo, C., Goedhals, D., Abrahams, S., Laguda-Akingba, O., Alisoltani-Dehkordi, A., Godzik, A., Wibmer, C.K., Sewell, B.T., Lourenço, J., Alcantara, L.C.J., Pond, S.L.K., Weaver, S., Martin, D., Lessells, R.J., Bhiman, J.N., Williamson, C. and de Oliveira, T. 2020. Emergence and rapid spread of a new severe acute respiratory syndrome-related coronavirus 2 (SARS-CoV-2) lineage with multiple spike mutations in South Africa. medRxiv 10.1101/2020.12.21.20248640. https://doi.org/10.1101/2020.12.21.20248640.

46. Volz, E., Hill, V., McCrone, J.T., Price, A., Jorgensen, D., O’Toole, A., Southgate, J., Johnson, R., Jackson, B., Nascimento, F.F., Rey, S.M., Nicholls, S.M., Colquhoun, R.M., da Silva Filipe, A., Shepherd, J., Pascall, D.J., Shah, R., Jesudason, N., Li, K., Jarrett, R., Pacchiarini, N., Bull, M., Geidelberg, L., Siveroni, I., Consortium, C.-U., Goodfellow, I., Loman, N.J., Pybus, O.G., Robertson, D.L., Thomson, E.C., Rambaut, A. and Connor, T.R. 2021. Evaluating the Effects of SARS-CoV-2 Spike Mutation D614G on Transmissibility and Pathogenicity. Cell 184(1), 64–75 e11.https://doi.org/10.1016/j.cell.2020.11.020.

47. Wang, H., Li, X., Li, T., Zhang, S., Wang, L., Wu, X. and Liu, J. 2020. The genetic sequence, origin, and diagnosis of SARS-CoV-2. Eur J Clin Microbiol Infect Dis 39(9), 1629–1635.https://doi.org/10.1007/s10096-020-03899-4.

48. Westhaus, S., Weber, F.A., Schiwy, S., Linnemann, V., Brinkmann, M., Widera, M., Greve, C., Janke, A., Hollert, H., Wintgens, T. and Ciesek, S. 2021. Detection of SARS-CoV-2 in raw and treated wastewater in Germany - Suitability for COVID-19 surveillance and potential transmission risks. Sci Total Environ 751, 141750.https://doi.org/10.1016/j.scitotenv.2020.141750.

49. WHO 2020a World Health Organisation - SARS-CoV-2 assay. https://www.who.int/docs/default-source/coronaviruse/whoinhouseassays.pdf?sfvrsn=de3a76aa_2

50. WHO 2020b World Health Organisation - Serology in the context of COVID-19. https://www.who.int/emergencies/diseases/novel-coronavirus-2019/serology-in-the-context-of-covid-19

51. Wu, F., Zhang, J., Xiao, A., Gu, X., Lee, W.L., Armas, F., Kauffman, K., Hanage, W., Matus, M., Ghaeli, N., Endo, N., Duvallet, C., Poyet, M., Moniz, K., Washburne, A.D., Erickson, T.B., Chai, P.R., Thompson, J. and Alm, E.J. 2020. SARS-CoV-2 Titers in Wastewater Are Higher than Expected from Clinically Confirmed Cases. mSystems 5(4), 00614–00620.https://doi.org/10.1128/mSystems.00614-20.

52. Wurtzer, S., Marechal, V., Mouchel, J.M., Maday, Y., Teyssou, R., Richard, E., Almayrac, J.L. and Moulin, L. 2020. Evaluation of lockdown effect on SARS-CoV-2 dynamics through viral genome quantification in waste water, Greater Paris, France, 5 March to 23 April 2020. Euro Surveill 25(50), 2000776.https://doi.org/10.2807/1560-7917.ES.2020.25.50.2000776.

53. Xing, Y.H., Ni, W., Wu, Q., Li, W.J., Li, G.J., Wang, W.D., Tong, J.N., Song, X.F., Wing-Kin Wong, G. and Xing, Q.S. 2020. Prolonged viral shedding in feces of pediatric patients with coronavirus disease 2019. J Microbiol Immunol Infect 53(3), 473–480.https://doi.org/10.1016/j.jmii.2020.03.021.

54. Yan, Y., Shin, W.I., Pang, Y.X., Meng, Y., Lai, J., You, C., Zhao, H., Lester, E., Wu, T. and Pang, C.H. 2020. The First 75 Days of Novel Coronavirus (SARS-CoV-2) Outbreak: Recent Advances, Prevention, and Treatment. Int J Environ Res Public Health 17(7).https://doi.org/10.3390/ijerph17072323.

55. Yue, J.C. and Clayton, M.K. 2005. A Similarity Measure Based on Species Proportions. Communications in Statistics - Theory and Methods 34(11), 2123–2131.https://doi.org/10.1080/sta-200066418.

56. Zhang, D., Ling, H., Huang, X., Li, J., Li, W., Yi, C., Zhang, T., Jiang, Y., He, Y., Deng, S., Zhang, X., Wang, X., Liu, Y., Li, G. and Qu, J. 2020. Potential spreading risks and disinfection challenges of medical wastewater by the presence of Severe Acute Respiratory Syndrome Coronavirus 2 (SARS-CoV-2) viral RNA in septic tanks of Fangcang Hospital. Sci Total Environ 741, 140445.https://doi.org/10.1016/j.scitotenv.2020.140445.

57. Zhang, Y.Z. and Holmes, E.C. 2020. A Genomic Perspective on the Origin and Emergence of SARS-CoV-2. Cell 181(2), 223–227.https://doi.org/10.1016/j.cell.2020.03.035.

