## Supplementary Figure 1 for "High-throughput sequencing of SARS-CoV-2 in wastewater provides insights into circulating variants"

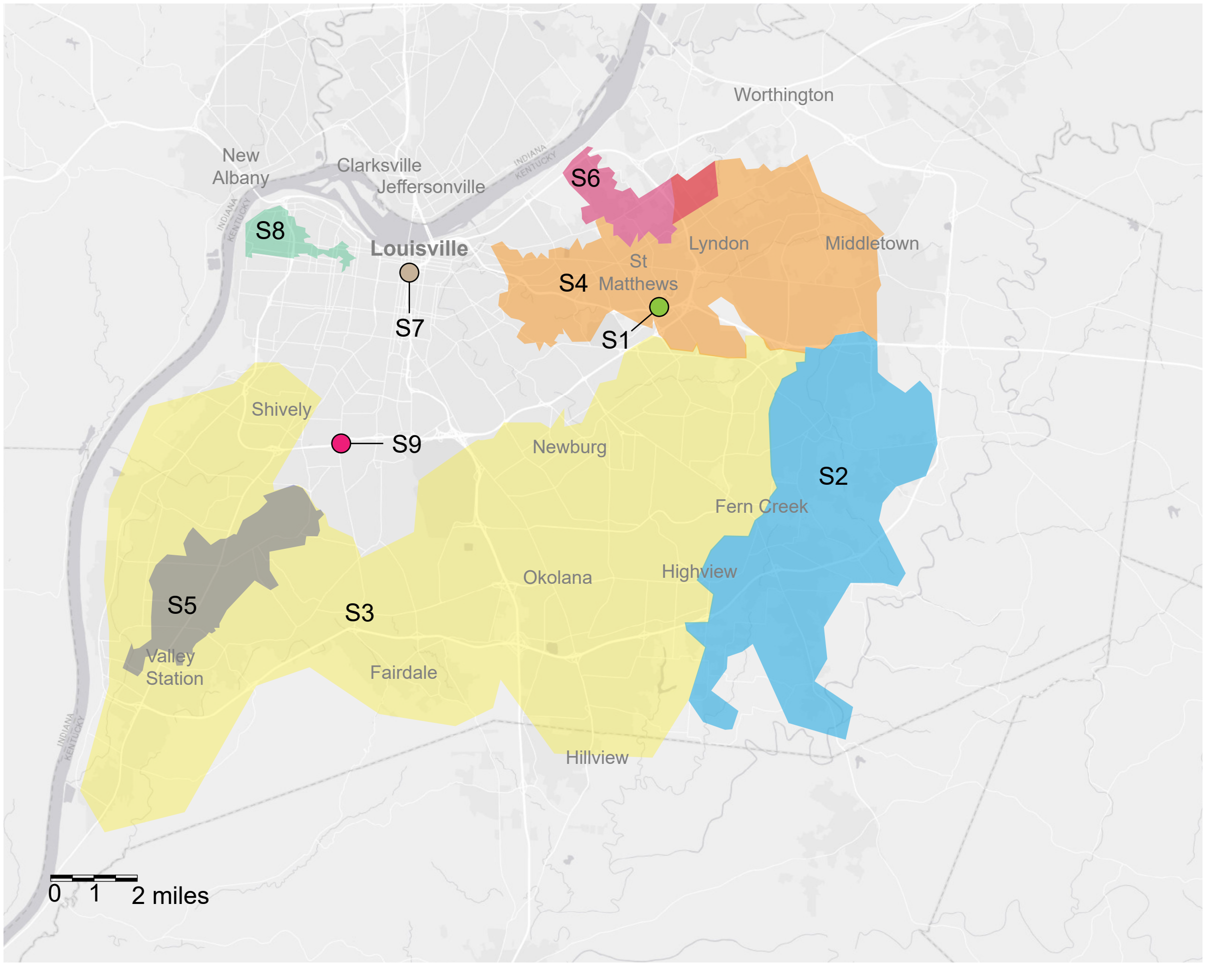


Supplementary Figure 1: Wastewater sampling catchments in Louisville (Kentucky), Sites 1 and 7 represent collection sites of hospitals and Site 9 is a sewer district facility.
